# “What do you mean I can’t have a doctor? This is Canada!” – A Qualitative Study of the Myriad Consequences for Unattached Patients Awaiting Primary Care Attachment

**DOI:** 10.1101/2021.07.07.21260143

**Authors:** Emily Gard Marshall, Sara Wuite, Beverley Lawson, Melissa K. Andrew, Lynn Edwards, Adrian MacKenzie, Ana Correa Woodrow, Sarah Peddle

**Affiliations:** Department of Family Medicine, Primary Care Research Unit, Dalhousie University, Halifax, Canada; Public Health, Nova Scotia Health, Halifax, Canada; Department of Medicine (Geriatrics), Dalhousie University, Halifax, Canada; Primary Health Care & Chronic Disease Management, Nova Scotia Health, Halifax, Canada; Research and Innovation, Nova Scotia Health, Halifax, Canada; Patient Partner, Halifax, Canada

**Keywords:** Primary health care, Primary Care, Access, Family practice, Waiting lists, Continuity of patient care

## Abstract

**Background:** Patient access to primary healthcare (PHC) is the foundation of a strong healthcare system and healthy populations. Attachment to a regular PHC provider, a key to healthcare access, has seen a decline in some jurisdictions. This study explored the consequences of unattachment from a patient perspective, an under-studied phenomenon to date.

**Methods:** A realist-informed qualitative study was conducted with unattached patients in Nova Scotia, Canada. Semi-structured interviews with nine participants were conducted and transcribed for analysis. The framework method was used to carry out analysis, which was guided by Donabedian’s model of assessing healthcare access and quality.

**Results:** Five key findings were noted in this study: 1) Participants experienced a range of consequences from not having a regular PHC provider. Participants used creative strategies to 2) attempt to gain attachment to a regular PHC provider, and, to 3) address their health needs in the absence of a regular PHC provider. 4) Participants experienced negative feelings about themselves and the healthcare system, and 5) stress related to the consequences and added work of being unattached and lost care.

**Conclusions:** Unattached patients experienced a burden of care related to lost care and managing their own health and related information, due to the download of medical record management and system navigation to them. These findings may underestimate the consequences for further at-risk populations who would not have been included in our recruitment. This may result in poorer health outcomes, which could be mitigated by interventions at the structural level, such as enhanced centralized waitlists to promote attachment. Such waitlists may benefit from a triage approach to appropriately attach patients based on need.

## Background

Patient access to primary healthcare (PHC) is the cornerstone of a strong healthcare care system; patient attachment to a regular PHC provider (a family physician or nurse practitioner) helps prevent illness and death (1). At the health system level, it is associated with better population health outcomes, lower overall costs of care, and reduced health disparities across socioeconomic status (2). In Canada, primary care is the first line of contact with the healthcare system, through which patients access diagnostics and further specialist care. For patients, attachment is related to more timely treatment (3), better preventive care and chronic disease management (4,5), lower likelihood of unmet health needs (6,7), and ultimately, better health outcomes (1,2). However, some populations face challenges to accessing care and experience lower attachment rates (7), such as recent immigrants (8), LGBTQ+ populations (9), and those experiencing homelessness (10).

Historically, population-level attachment to a PHC provider has been high in Canada. The percentage of Canadians with a regular PHC provider grew significantly from 83.2% in 2015 to 85.5% in 2019 (11). However, in Nova Scotia, a province with an aging population and high burden of chronic illness, attachment instead dropped significantly from 88.7% to 85.6% throughout that same time period (11). Among those Nova Scotians without a PHC provider, 41% reported that they previously had a provider who had left or retired (11). Because of this, there has been widespread public scrutiny regarding the lack of PHC providers in the province (12), an issue which has grown in the wake of the COVID-19 pandemic (13). Provincial governments across Canada are responsible for healthcare operations and delivery, and recognize the importance of PHC attachment. As of May 2021 seven Canadian provinces, including Nova Scotia, have implemented centralized waitlists for their unattached patients in an attempt to streamline attachment processes and increase attachment rates (14).

As patients move through phases of becoming unattached and back to attachment, such as while they are on a centralized waitlist, they may use creative strategies to access care and to find a new PHC provider (15,16). However, little is known about what the consequences of unattachment are from a patient perspective or about the strategies that unattached patients use to meet their health needs and access care. The purpose of this study is to inform health system strategies for mitigating negative consequences of unattachment and for increasing attachment by understanding patient-defined impacts of unattachment and the strategies they use to access PHC. The study was guided by the following research questions: a) What are the lived experiences of unattached patients in relation to their health, healthcare needs, and attempts to find a regular PHC provider; and b) How are unattached patients’ experiences related to the social determinants of health and social vulnerability?

## Methods

We conducted a qualitative study informed by a realist orientation, which focuses on the experiences and realities of participants (17). This methodological approach is appropriate as the topic of unattachment in PHC is novel in the literature, with only two qualitative studies in chronically-ill subpopulations found to date (15,16). The perspectives of unattached patients, especially in this geographic region, are unknown and require an exploratory approach. Our analysis was informed by Donabedian’s model of assessing healthcare access and quality, focused on structure, process, and outcomes (18), and operationalized using the framework method of qualitative analysis as outlined below (19).

Our study was approved by the Nova Scotia Health Authority Research Ethics Board (File #1022763). We recruited unattached patients using invitational letters to patients on the Nova Scotia Need a family Practice Registry. To maintain the confidentiality of people on the registry, letters were sent by the registry custodians on behalf of the study team. We also advertised on social media and local online marketplaces. For all recruitment methods, potential participants were instructed to contact the study team if they were interested in participating. To be considered for inclusion, potential participants had to be over 18, speak English, reside in Nova Scotia, and not have a regular family physician or nurse practitioner. After receiving expressions of interest via email from potential participants, we conducted pre-screening phone calls and iteratively selected participants using purposive sampling based on geography, age, gender, and healthcare needs (20,21). We continued to invite participants until saturation was reached (22).

A trained qualitative researcher conducted face-to-face interviews in locations that were convenient for each participant. The interview guide was developed by the study team, based on unattached patient team member experience. Interviews lasted approximately one hour, were digitally recorded, and transcribed verbatim.

Interview data were coded inductively and deductively. Initial transcript coding was conducted by four team members, including the interviewer and principal investigator; final coding of all transcripts was conducted by the interviewer. Initial coding categories were developed based on the experience of unattached patient team members. Additional codes were added inductively from the transcript data. The final coding and thematic framework structure were developed iteratively and collaboratively among four team members, including an unattached patient team member. All analysis was conducted in NVivo 11 (QSR International).

## Results

We interviewed nine participants with varied socio-demographic representation (Table 1).

**Table 1:** Socio-demographic characteristics of unattached patient study participants

|  | **Number of Participants (%)** |
| --- | --- |
| **Gender** |  |
| Male | 3 (33%) |
| Female | 6 (66%) |
| **Age (years)** |  |
| 18-39 | 2 (22%) |
| 30-49 | 1 (11%) |
| 40-49 | 1 (11%) |
| 50-59 | 1 (11%) |
| 60-69 | 4 (44%) |
| **Health Status (self-report)** |  |
| Very good | 7 (77%) |
| Good | 2 (22%) |
| **Visible Minority** |  |
| Yes | 2 (22%) |
| No | 7 (77%) |
| **Highest Education** |  |
| High School | 1 (11%) |
| Trade Diploma | 2 (22%) |
| University Certificate or College Diploma | 3 (33%) |
| Bachelor's | 3 (33%) |
| **Employment Status** |  |
| Employed | 5 (55%) |
| Self-Employed | 1 (11%) |
| Other | 3 (33%) |
| **Household Income (\$CAD)** | |
| 10,000-19,000 | 1 (11%) |
| 20,000-29,000 | 2 (22%) |
| 30,000-39,000 | 0 |
| 40,000-49,000 | 0 |
| 50,000-59,000 | 0 |
| 61,000-69,000 | 0 |
| 70,000-79,000 | 2 (22%) |
| 80,000-89,000 | 1 (11%) |
| 90,000-99,000 | 1 (11%) |
| 100,000-149,000 | 1 (11%) |
| 150,000+ | 1 (11%) |

Participants became unattached through both patient- and provider-initiated mechanisms. Most commonly, participant moved either within the province or from outside of the province. In some cases, a previous PHC provider retired or moved out of province. In one instance, a participant had a previous PHC provider that may have been available to them after moving back to Nova Scotia, but they were uncomfortable receiving care from them and so chose to look for a new regular PHC provider. No participants in our sample were “fired”(23) from their most recent PHC provider before participating in the study.

Guided by framework analysis (19), five main findings from interviews with unattached patients were uncovered and are described in more detail below: 1) Unattached patients experience a range of consequences from not having a regular PHC provider. Unattached patients use creative strategies to 2) attempt to gain attachment to a regular PHC provider and to 3) address their health needs in the absence of a regular PHC provider. Unanimously, 4) participants experienced stress related to the consequences and added work of being unattached and 5) had negative feelings and sense of abandonment from not having a regular PHC provider.

### Consequences of Unattachment

Participants experienced a range of consequences from being unattached related to the burden of care, lost care, and health impacts (Figure 1).

**Figure 1:**
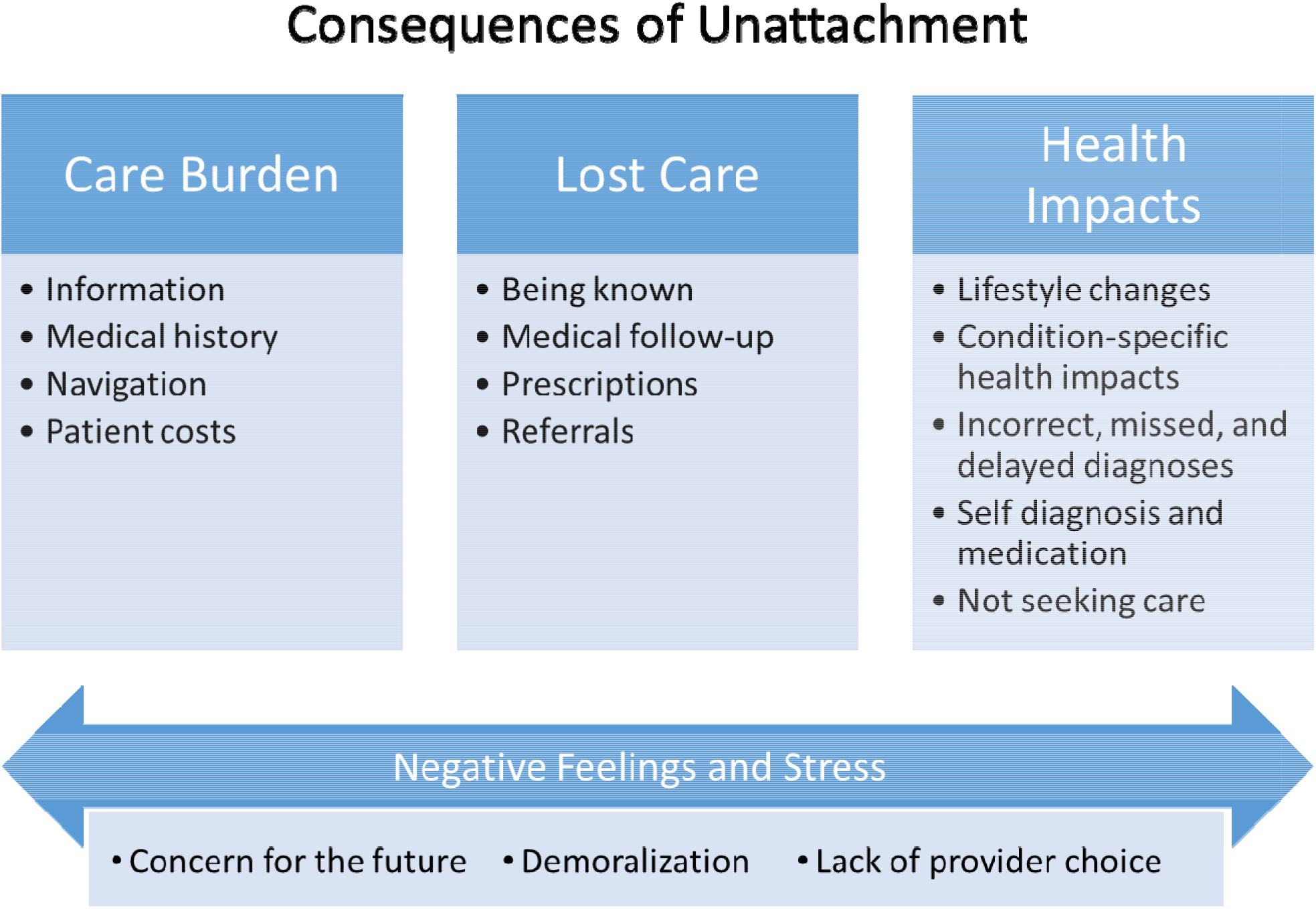
Themes related to consequences of unattachment.

#### Care Burden

All participants experienced an increased burden of managing their own healthcare in the absence of a regular PHC provider. Participants experienced increased burden compared to when they were attached for finding and managing information, managing their own medical history, navigating health and other service systems, and in some cases, patient costs including time and increased travel distances for care.

Participants look to their PHC providers as experts in medicine and in the healthcare system. Without a regular PHC provider, several participants looked to other sources for information that may have some healthcare expertise, such as the internet or family and friends.

Three participants expressed that they specifically felt the loss of an expert who could give concrete answers and to help process information into something actionable.

> *“Kind of that hub where, you know, all the information goes in and somebody needs to kind of figure out what to do with it” – Participant 8*

All participants discussed the burden of having to manage their own medical record while unattached. This manifested as needing to communicate their medical history to interim providers, managing new healthcare information and instructions that would typically be held in a medical record under the custodianship of a regular PHC provider, and extra work to track and access their existing medical records from previous PHC providers. Participants expressed that they felt stressed by these added responsibilities and worried that they may not be able to carry them out as well as a regular PHC provider. One participant took detailed notes in order to maintain their medical history while accessing walk-in clinics; another paid to have a print copy of her whole medical history up until they became unattached.

> *“…I did a lot of talking to bring the [walk-in clinic] doctor up to speed … what happens now if I got to somebody that I don’t know and doesn’t know me or my history, and I have 15 minutes or whatever, and they’re absolutely overloaded with patients anyway?” – Participant 1*
>
> *“Because when I got to the walk-in, and I tell them I had a surgery once but they don’t really know much about it, and I can’t tell them a whole lot about it … Like, I am not very good at, you know, explaining things when I am on the spot*.*”-Participant 4*

At the intersection of the loss of a “home” for their medical records and an increased burden of managing healthcare information, our study participants expressed the consequence of having to navigate the healthcare system alone or with dis-jointed advice. Some participants expressed confusion about how long is normal to wait for a follow-up from a referral or who is even responsible for initiating that follow-up. Others felt challenged by having to navigate different care providers and services without the central “hub” of a PHC provider to help coordinate their healthcare around a particular health issue.

> *“You’re just sort of roaming around a maze, you know, with drones at each station*.*” –Participant 3*

Although healthcare in Canada, including PHC, walk-in clinics, and hospital care, is funded through taxes and has no user access fees, some services, such as purchase of most prescription drugs and services of some allied health professionals, incur fees unless covered by an insurance plan. Participants in this study faced increased financial and time costs related to not having a regular PHC provider. Most commonly, participants sought over the counter medications, where if they had a regular PHC provider, they would have sought a prescription (or other treatment avenue) that would then be covered through their medical insurance. Four participants had costs related to accessing their medical records or sending medical information to their insurance companies. Several participants also expressed costs related to travel, including going to walk-in clinics or other healthcare services that are further than they would typically travel; paying for taxis or in one case, an ambulance; and time away from work having to wait in emergency rooms and walk-in clinics that do not offer pre-booked appointment times.

#### Lost Care

All patients had trouble accessing certain kinds of care and felt they “lost” some care due to not having a regular PHC provider. There was a strong theme of the benefit of “being known” to a regular PHC provider, which supported participant’s comfort in accessing care and providers’ ability to provide continuous care. Without a regular PHC provider, participants did not feel “known” by the interim providers they saw. This, grouped with the unavailability of their whole medical history and bouncing between various interim care providers, resulted in feeling that pieces of their care got lost.

> *“Having to tell your story over and over and over again to …and also I think for a physician, they may be missing certain links. I think the impact has been enormous, actually” – Participant 5*
>
> *“The first time [with an emerging medical issue] I bounced from doctor to doctor each visit … History matters. It matters very much. Yeah, you know your doctor and your doctor knows you” – Participant 2*

All participants faced challenges, sometimes insurmountable, in accessing either medical follow up from a visit to an interim provider, prescriptions, or referrals to specialists. In particular, getting prescriptions for controlled medications – such as those for ADHD, sleeping, and other mental illnesses – posed serious problems at walk-in clinics, even if the participant had been prescribed the medication they were seeking previously. Similarly, some participants had trouble accessing referrals to specialists or for testing, and then if a referral was made, were challenged by a lack of a “home” to manage follow-up, such as receiving test results and next steps. For example, one patient struggled to have an issue diagnosed via walk-in clinics but received one upon an emergency room visit for acute pain related to the issue. From the emergency room, they were referred to a surgeon for treatment. The participant was unsure of how long they should wait to hear from the surgeon; eventually, it became clear that the referral either was not made or got lost. They then struggled to gain another referral from interim PHC sources.

> *“I mean, I didn’t know if I could have called the emergency room to ask who I had been referred to. It didn’t seem like that was a place I could sort of go back to to follow up. So, you just sort of feel a little bit sort of forgotten and kind of lost in the system. Like I think I got lost in the system a couple of times, and I didn’t really know how to follow up on it or just what to do next*.*” – Participant 9*

#### Health Impacts

When asked directly if being unattached had direct impacts on their health, most participants responded that it did not. However, when probed further about specific experiences, health impacts emerged, including making lifestyle changes to improve their health, condition-specific changes in health, incorrect, missed, or delayed diagnoses, self-diagnosis and medication, and not seeking care.

Three participants made proactive lifestyle changes in an attempt to reduce their healthcare needs and to manage existing symptoms. Lifestyle changes included diet, exercise, being more cautious, and occupational health changes (e.g., shoe insoles for walking, physiotherapy-inspired exercises for office work).

> *“Even walking down stairs, like even when I’m moving around, I’m thinking just take a second and think about what you’re doing…I’m trying to keep myself from need to see [a doctor]*.*” –Participant 3*

Six participants experienced some deterioration in their health or sustained symptoms, related to a specific health issue that they each experienced, that they attributed to not having a PHC provider.

> *“…kind of waiting for these pain flare-ups to happen again because it would sort of be unexpected flare-ups with terrible pain*.*” – Participant 9*
>
> *“It’s just been kind of the same things having to deal with several years or longer. For a long time at this point. So, it just kind of ends up becoming the background noise of my life” – Participant 7*

Seven participants felt that they had diagnoses that were either incorrect, delayed, or missed altogether due to the discontinuity of PHC and the absence of having a complete medical record available for interim care without a regular PHC provider. For example, one participant had a persistent issue for which they sought care at a walk-in clinic several times and then the emergency room. They eventually had to call an ambulance and seek emergency care for the issue. Another participant spent five months with debilitating symptoms before receiving a diagnosis that was ultimately picked up with routine bloodwork, which had never been ordered by the walk-in doctors she visited previously.

In the absence of a regular PHC provider, several participants reported self-diagnosing and treating their health conditions, despite their perception that this was not “the right” thing to do. Relatedly, some participants reported not seeking any care even when they felt it was warranted and would have if they had a regular PHC provider. In both cases, participants felt either the barriers of finding appropriate “professional” healthcare were insurmountable (e.g., waiting at a walk-in or in emergency), or, that their healthcare needs were not severe enough to warrant discontinuous walk-in care or emergency care.

> *“So, you don’t feel you should go in unless you’ve really got something serious wrong with you. So yeah, you do self-medicate” – Participant 2*
>
> *“And I guess the biggest difference is we sort of feel held back from dealing with certain issues because it is not convenient to be able to access the care. So maybe we would have gone to the doctor for less of a reason” – Participant 6*

### Strategies for Care: Attachment Strategies

#### Unattached Patients’ Registry

Most participants were registered on the provincial unattached patient registry. Only one of the participants attached through the registry at the time of their interview, and none of the participants on the registry held much hope that they would find a provider through this strategy.

> *“And I always kind of figured that I wouldn’t be on the list for very long … And so, I figured it was just a matter of time before they called me or contacted me at all. But no, they never did. They still haven’t*.*” – Participant 7*
>
> *“But we put ourselves on the list with the government for all the [expletive] good that’s going to do” – Participant 3*

#### Interim PHC Provider

A common strategy was to gain or approximate attachment through an interim care provider. Participants would either try to attach to the regular practice of the doctors that they saw in walk-in clinics or rural emergency rooms, or they would attempt to return to the same walk-in clinic doctor each time they visited. In one case, a walk-in doctor agreed to see a patient continuously until a particular health issue was resolved.

#### Cold-Calling PHC Practices

Another common practice was to cold-call PHC offices to see if any of the providers were taking new patients. Six of the participants tried this approach, but half have since stopped due to feelings of discouragement. One participant had a dedicated approach of calling all the practices in a geographical area that feels acceptable to them every two months. This method elicited a lot of frustration among participants, but also empathy for the receptionists who receive their calls, whom participants described as generally pleasant and sympathetic. No participants found a new provider with this approach.

> *“I’ve stopped [calling clinics] because it’s pointless. You know, I’m well aware of how bad it is now. So, I’ve stopped trying” – Participant 6*

#### Personal Networks

Seven participants used their personal networks to try to find a new regular PHC provider. Participants’ approaches ranged from asking friends and family if they knew of any providers taking new patients, to requesting that they ask their providers directly (e.g., a mother asked if her physician would take her unattached son), to asking directly among PHC providers that they knew personally. There were mixed approaches to using social media – some participants used all networks available to them, while some did not feel comfortable “broadcasting” their healthcare issues in such a public way. Two participants indicated that they used public and personal political avenues to try to inspire broader change or simply understand the larger issue of unattachment to be better able to navigate the system to find a provider for themselves.

> *“You know, the only way you’re going to find a doctor in NS is if you know somebody or if somebody happens to move into a new practice*.*” –Participant 3*

#### Expanding Geographic Area

Regardless of the active approach to finding a new PHC provider, four participants expressed a willingness to attach to a provider in a geographic area outside of what they consider convenient. Some participants expressed frustration that you could not indicate willingness to travel on the unattached patients’ registry.

#### Giving Up

Finally, most participants simply gave up actively looking for a regular PHC provider.

> *“…there is no doctor to be found. You know, we start paying attention to the news and … you know. And it was the start realization that, you know what, it’s a systemic problem in this province” – Participant 6*

### Accessing Care While Unattached

Similar to not immediately identifying health impacts related to being unattached, participants frequently answered that they did not have any ongoing healthcare needs while unattached. However, as participants’ narratives unfolded, it became clear that most had either an emergent healthcare need or a controlled chronic condition during their unattachment. While participants did not always identify a specific need in the time that they were unattached, most expressed concern for the future, when things “get worse” or “come up.”

#### Walk-in Clinics

Walk-in clinics were the primary source of PHC for participants while unattached. Some participants could access walk-in hours in the family practice clinic they used to attend, others used dedicated walk-in clinics that provide no continuous care. While some were able to use the same walk-in clinic repeatedly to approximate attachment, most found them to be frustrating due to wait times, feeling rushed in the appointment, reluctance of walk-in doctors to order tests, write referrals, or write certain prescriptions, having to be the reporter of their own medical history, and the perceived incompetence of some walk-in physicians. While some participants were thankful to have access to some form of care, most felt walk-ins were an inadequate replacement for a regular PHC provider.

> *“But having been to a couple of walk-in clinics for various things, like, you know, a sinus infection for my daughter or strep throat for my son or whatever, and having to deal with these people at these walk-in clinics was enough to turn me off and say, you know what, it’s not worth dealing with*.*” – Participant 6*

#### Emergency Care

Overall, participants were reluctant to use emergency care unless they felt their issue was truly an emergency. In some cases, participants did use emergency care when they felt it was warranted, but that it could have been avoided if they had a regular PHC provider to address their issue before it escalated.

#### Specialists and Other Healthcare Professionals

Participants used a variety of healthcare providers that were available to them. Two participants had access to specialists for other ongoing health issues and had those specialists address issues outside of their area of expertise that would typically be addressed by a PHC provider. Pharmacists were accessed by seven participants in lieu of a PHC provider, primarily for preventative health services like flu shots. Participants also access self-referral walk-in clinics, methadone clinic physicians, chiropractors, and women’s health clinics for issues that would be appropriate for PHC.

### Stress of Managing Unattachment and Concerns for Future Health

Overwhelmingly, due to the consequences and the increased work for accessing care, all participants experienced stress related to being unattached. Most participants expressed concern for the future, related to accessing care for either an exacerbation or recurrence of an existing health condition or for a new issue, often related to aging. This stress was especially pronounced among participants who had chronic health conditions that would benefit from a PHC provider’s involvement. Several participants also expressed frustration and worry about the perceived lack of choice of providers – both at walk-in clinics and with a potential new regular PHC provider.

> *“It probably wouldn’t bother me a lot but I’ve been through cancer treatment. So I’m in remission. But I have ongoing care from that experience. But what I don’t have is somebody [to care for me] if something else comes up even related to my cancer*.*” - Participant 1*
>
> *“I am fortunate right now that things are kind of stabilized and I am healthy. But you know, if that changes, I’m not sure what I would do*.*” – Participant 9*
>
> *“Health and stress go together in my life” – Participant 3*

### Negative Feelings and Sense of Abandonment due to Unattachment

Finally, several participants expressed an unexpected feeling of abandonment or betrayal by the healthcare system due to not having a regular PHC provider. Participants expressed feeling demoralized and “like a beggar” in the process of seeking a new PHC provider. They also expressed a desire to have access to a regular PHC provider and described the sense of comfort and reassurance they would feel if they could become attached.

> *“Yeah, you’re not expecting it. It’s kind of shocking. And then to realize there are no doctors. What do you mean I can’t have a doctor? This is Canada! We all have medical care, right? No, we don’t. It was startling*.*” – Participant 2*
>
> *“The biggest thing is not having that level of comfort. You know, that it’s not… It’s a bit of a security blanket to know that you have a family doctor*.*” - Participant 6*

## Discussion

Unattached patients experience a range of consequences related to not having a regular PHC provider. They also use a variety of creative strategies to seek attachment and to address their healthcare needs. Our study found that unattached patients experience a burden of care related to managing their own medical records and worry about having to be the reporter of their own medical history, especially when it is with an unfamiliar provider. This is consistent with the findings of another qualitative study in British Columbia that found similar themes related to medical records and a desire for a trusting relationship with a regular PHC provider (16). Interestingly, patients were able to keenly identify the stress associated with not having a regular PHC provider, but until prompted about specific experiences, were hesitant to express specific health consequences or even healthcare needs. While participants were creative in their efforts to obtain interim care, it was clear that many still had healthcare needs that went unmet. Indeed, not having a regular PHC provider is a significant and strong predictor of having unmet healthcare needs (7).

Another small study of unattached women with chronic illness identified similar strategies to accessing interim care and attachment, including relying on specialists until their healthcare needs exceeded the specialists’ scope, and using walk-in clinics; this study also identified some of the same frustrations with walk-in clinics, including wait times, inability to book an appointment, and seeing different PHC providers each time (15). This study also found similar strategies to gain attachment, including calling practices and a willingness to expand geographical scope.

Building on these two qualitative studies on unattachment, our study adds a deeper understanding of the impact of these added care responsibilities for a general population of unattached patients, ranging from self-diagnosis and treatment through to giving up on both seeking care and looking for a new PHC provider, improving health behaviours to avoid needing care, and a negative feeling of abandonment from a health care system though thought they could always rely on.

While we reached data saturation, our study is still limited by a small number of volunteer participants within one province in a country with provincially delivered healthcare. Our recruitment methods were also not designed to reach marginalized populations, members of cultural and language minorities, socially isolated and/or vulnerable people, older adults, those with limited access to technology, those with severe health conditions, or those with cognitive impairment who are likely to experience amplified or different consequences of unattachment, and for whom the compensating strategies to meet health care needs described by our participants may not be feasible. However, recruited participants who were relatively empowered and particularly motivated to share their experiences, our purposive sampling method allowed for participant selection across several socio-demographic variables and unattachment experiences, highlighting the pervasiveness of negative consequences of unattachment across sociodemographic characteristics that are not limited to those with more challenging circumstances.

## Conclusions

Our study demonstrates the impact of unattachment and can be understood using the Donabedian model of healthcare access of structure, process, and outcomes (18). While unattached, much of the “process” of healthcare, including medical record management and system navigation, is downloaded from PHC providers onto patients, which may contribute to poorer health and social outcomes. Similarly, patients are responsible for seeking new attachment via centralized wait lists and other means, as well as their care while unattached. Patients may also have differential capacity to pursue attachment and may face discrimination in that process (24). Hence, first-come-first-served centralized waitlists may be an insufficient approach that has the potential to be unfair and result in inequitable care and re-attachment along socio-demographic axes. As such, unattached patients may benefit from interventions at the “structure” level, such as health human resource interventions to ensure an adequate PHC workforce and patient registries/wait lists to facilitate patient attachment to PHC.

Given that patients with ongoing health needs cited greater stress related to not having a PHC provider relative to patients without a regular PHC provider, our study suggests that patient registries and waitlists may best support unattached patients by taking a triaged approach to prioritize attachment for patients with greater needs, rather than using a purely first-come-first-served, time-based algorithm. The more severe health impacts for participants that were managing an acute healthcare need suggest that these people (and the health system as a whole) may have benefited from more efficient attachment than those who did not have any severe chronic or acute care concerns. Participants’ “concern for the future” when not perceiving an immediate healthcare need and their reluctance to use emergency rooms unless absolutely necessary suggests there is an openness among unattached patients to ranking need to shepherd efficient and effective attachment. Triaged registries exist in British Columbia, Ontario, and Quebec (14), where they have demonstrated effectiveness at prioritizing vulnerable patients (25), with other structural factors promoting even stronger attachment for vulnerable patients (26).

Last, but not least, it is critical to consider how a potentially growing lack of confidence in the health care system’s ability to provide access to having a regular primary care provider may impact support for Canada’s decentralized, universal, publicly funded health system. Next steps include sharing these findings with our provincial government, health authority, and patient partner stakeholders who are active members of the study team. Future work could use survey methods to observe the spread and scale of the concerns and consequences identified by our research participants, and the potential impact of the COVID-19 pandemic on access to primary care attachment.

## Data Availability

Due to the identifiable nature of raw data (interview transcripts), they are not publicly available. Requests for additional information about study data or results can be forwarded to the corresponding author (EGM).

## Declarations / Acknowledgements

### Ethics approval and consent to participate

This study was approved by the Nova Scotia Health Research Ethics Board (#1022374). The informed consent discussion was conducted by a trained master-level interviewer, and participants provided written informed consent.

### Consent for publication

Not applicable.

### Availability of data and materials

The data that supports the findings of this study are not publicly available due to the identifiable nature of raw data (interview transcripts). Requests for additional information about study data or results can be forwarded to the corresponding author (EGM).

### Authors’ contributions

EGM conceptualized the study, designed the methodology, established protocol, analyzed and interpreted data, provided software, writing the manuscript, supervision of data collection and analysis, funding acquisition. SW collected, analyzed and interpreted data, manuscript writing and editing. BL supported funding application, study development, analysis review, writing-reviewing, and editing. MKA supported funding application, study development, analysis review, writing-reviewing, and editing. LE supported funding application, study development, writing-reviewing, and editing. AM supported funding application, study development, analysis review, writing-reviewing, and editing. ACW supported funding application, study development, analysis review, writing-review, active Patient Partner for this study. SP supported funding application, study development, analysis review, writing-review and editing, active Patient Partner for this study.

## Acknowledgements

We gratefully acknowledge our study participants who generously shared their time and lived experiences as unattached patients.

